# Work Fatigue in Phosphoric Acid Industry Workers: How Work shift and Sleep Quality Affect Them?

**DOI:** 10.1101/2023.05.21.23290287

**Authors:** Dayinta Annisa Syaiful, Indriati Paskarini, Tri Martiana, Angela Tesalonika Oktavera

## Abstract

**Introduction:** Manpower is a firm asset and it plays a role in determining the quality and quantity of items produced. Labour-intensive activities require a lot of energy. Work fatigue is one of the repercussions experienced by the workforce. This study aims to investigate the relationship between work shifts and sleep quality and subjective work fatigue at the phosphoric acid industry.

**Material and Method:** This study was conducted on all 44 workers using a cross-sectional technique. The PSQI questionnaire and the Subjective Feelings of Fatigue questionnaire was conducted to obtain the data. Spearman correlation and contingency coefficient test were used to analyse the data.

**Result:** There are 47.7% employees experienced moderate work fatigue. There was a strong relationship between work shift (p=0.637) and sleep quality (p=0.619) with work fatigue.

**Conclusion:** Special inspection at 2-4 am should be conducted to monitor the employees.

## BACKGROUND

Manpower is a company asset in conducting business in the industrial sector and has a role in determining the quality and quantity of goods to be produced. Activities carried out by labour are physical activities that drain a lot of energy. One of the consequences borne by the workforce is the emergence of work fatigue. Work fatigue is a decrease in work efficiency, performance, boredom arises and an increase in anxiety. Work fatigue contributes approximately 50% to the incidence of work accidents (Setyawati, 2007).

Fatigue is a mechanism in the body that aims to protect the body from further damage. The condition of the body can recover after the body experiences rest (Tarwaka, 2004). Work fatigue arises when the worker is no longer able to do his job. People who experience work fatigue are usually characterized by symptoms such as feeling sluggish, yawning, drowsiness, dizziness, difficulty thinking, lack of concentration, lack of alertness, poor perception and slowness, stiffness and awkwardness in movement, reduced desire to work, unable to stand. balance, tremors in the limbs, difficulty controlling attitude, and decreased physical and spiritual performance (Tarwaka, 2014).

Fatigue denotes different states, but the state of this leads to a reduction in work capacity and resilience body. There are two types of fatigue: muscle fatigue and general fatigue. Muscle fatigue is a tremor in the muscles, whereas general fatigue is characterized by a reduced ability to work. General fatigue is caused by monotonous work, work intensity, length of mental and physical work, environmental conditions, mental causes, nutritional status, health status, and workload (Suma’mur, 2009).

The International Labor Organization (ILO) states that the number of accidents in the workplace reaches more than 250 million annually and more than 160 million workers experience illness due to hazards in the workplace. Around 1.2 million workers die from accidents and illness at work. The results of a survey in the United States showed that 24 percent of all adults who came to the polyclinic suffered from fatigue (Setyawati, 2010).

Based on ILO data, in 2010 every year as many as 2 million workers died due to work accidents caused by fatigue. Based on this research, 32.8% of the 58,115 samples or about 18,828 samples suffered from work-related fatigue. Factors that affect fatigue include the intensity and duration of physical and mental work, monotomies, work climate, lighting, noise, responsibility, anxiety, conflict, illness, and nutrition (ILO, 2013).

## MATERIALS AND METHOD

This research is an observational research type because the data is obtained through a questionnaire in the field and without any treatment on the object under study. Based on the implementation time, this research is a cross sectional study because the variables are studied at one time. The population of this study is 44 workers in phosphoric acid plant and the sampling method used was total sampling. All workers in phosphoric acid plant were included in this study. The data obtained was among March until June 2021.). The study received ethics approval from the Faculty of Dentistry, Airlangga University, Surabaya (certificate number 275/HRECC.FODM/VI/2021)

The instrument used in this study was a questionnaire containing questionnaires on individual characteristics of respondents, work shifts, Pittsburgh Quality Sleep Index (PQSI) questionnaires, and subjective feelings of fatigue questionnaires from the Industrial Fatigue Research Committee (IFRC).

Validity and reliability tests were carried out on one variable, namely sleep quality. Validity and reliability tests were conducted based on research that also translated the Pittsburgh Sleep Quality Index (PSQI) questionnaire.

PSQI is a questionnaire that has been used in many studies with sleep quality parameters. The validity test uses the Pearson Product Moment correlation coefficient formulation. The result of the test is that the correlation level of r count 0.487 – 0.778 (r table value > 0.444) has the meaning of meeting the significant level. Reliability testing using Cronbach’s Alpha reliability coefficient formulation shows an alpha number of 0.841 which means that Cronbach’s alpha value > 0.6 indicates that the instrument is reliable (Fatmawati, 2013).

The data obtained were analyzed statistically by correlation test. Correlation test is used to determine the relationship between two variables. The variables tested are the dependent variable and the independent variable. This study uses the correlation test method, namely the Spearman correlation test and the contingency coefficient test (c). Spearman’s test was used for ordinal scale data analysis, while contingency coefficient test was used for nominal scale data analysis.

## RESULT

### Work shift in Phosphoric Acid Plant

Work shift is the arrangement of the working time of workers in groups work, assigned by the company. In this study, work shift is categorized into three groups, namely morning shift, afternoon shift, and night shift. The following shows the distribution of work shift

Based on the table 1, it can be seen that as many as 22 respondents or 50.0% of the total respondents worked on the morning shift. Meanwhile, 11 respondents or 25.0% of the total respondents worked on the afternoon shift, and 11 other respondents or 25.0% of the total respondents worked on the night shift.

**Table 1.** Distribution of Work shift in Phosphoric Acid Plant

| No. | Work shift | Amount (People) | Percentage (%) |
| --- | --- | --- | --- |
| 1. | Morning (07.00 – 15.00) | 22 | 50.00 |
| 2. | Afternoon (15.00 – 23.00) | 11 | 25.00 |
| 3. | Evening (23.00 – 07.00) | 11 | 25.00 |
| Total |  | 44 | 100 |

### Sleep Quality in Phosphoric Acid Plant

Sleep quality is a very complex matter, including assessment of sleep duration, sleep disturbances, sleep latency, daytime sleep dysfunction, sleep efficiency, sleep quality, and use of sleeping pills. Sleep quality is categorized into two groups, namely good sleep quality and poor sleep quality. Distribution of respondents based on sleep quality is described in the following table

Table 2 explained the distribution of sleep quality of workers at the phosphoric acid plant at PT. X. The distribution of the education level that dominates the poor sleep quality is 68.18% of the total research respondents.

**Table 2.** Distribution of Sleep Quality in Phosphoric Acid Plant

| No. | Sleep Quality | Amount (People) | Percentage (%) |
| --- | --- | --- | --- |
| 1. | Good | 14 | 31.82 |
| 2. | Bad | 30 | 68.18 |
| Total |  | 44 | 100 |

### Subjective Work Fatigue in Phosphoric Acid Plant

Work fatigue is a condition due to work carried out by workers in the form of feelings of fatigue and decreased ability to carry out an activity based on subjective methods felt by the workforce.

Based on table 3, it can be seen that most of the workers in the phosphoric acid plant at PT. X experienced a moderate level of fatigue, which was 47.7% of the total research respondents.

**Table 3.**
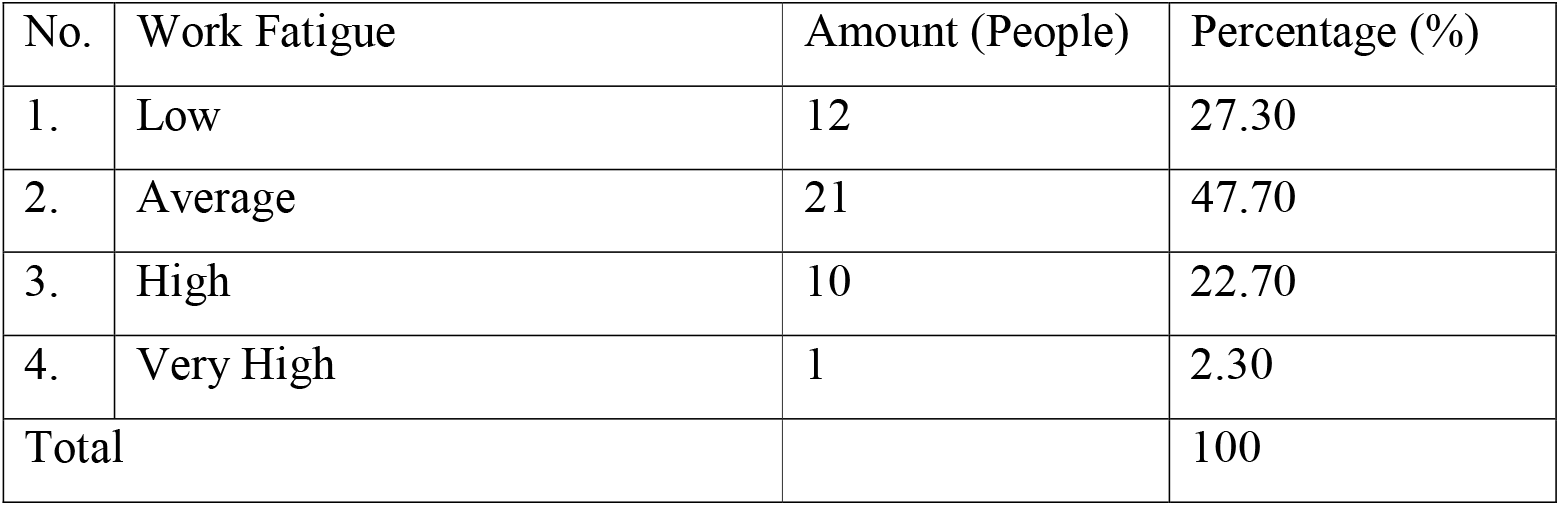
Distribution of Work Fatigue in Phosphoric Acid Plant

| No. | Work Fatigue | Amount (People) | Percentage (%) |
| --- | --- | --- | --- |
| 1. | Low | 12 | 27.30 |
| 2. | Average | 21 | 47.70 |
| 3. | High | 10 | 22.70 |
| 4. | Very High | 1 | 2.30 |
| Total |  |  | 100 |

### Association between Work shift and Work Fatigue in Phosphoric Acid Plant

The results obtained using cross tabulation between work shift with subjective work fatigue, the following data are obtained:

Based on table 4, it can be seen that respondents in the phosphoric acid plant with morning work shifts (07.00 – 15.00) the majority experienced moderate fatigue of 45.45%, while respondents with afternoon work shifts (15.00 – 23.00) experienced moderate fatigue of 81.82 %, and respondents with night shifts experienced high work fatigue of 72.72%. This shows that respondents with night shifts tend to experience high fatigue compared to morning shifts and afternoon shifts. From the results of statistical analysis, it was found that the correlation coefficient between the age variable and the work fatigue variable was 0.637, which means the strength of the relationship between work shifts and work fatigue is strong.

**Table 4.** Association of Work Fatigue based on Work Shift

| Work Shift | Work Fatigue |  |  |  |  |  |  |  | Total |  | Correlation Coefficient |
| --- | --- | --- | --- | --- | --- | --- | --- | --- | --- | --- | --- |
|  | Low |  | Average |  | High |  | Very High |  |  |  |  |
|  | N | % | N | % | N | % | N | % | N | % |  |
| Morning | 10 | 45.45 | 10 | 45.45 | 2 | 9.10 | 0 | 0 | 22 | 100 | 0.637 |
| Afternoon | 2 | 18.18 | 9 | 81.82 | 0 | 0 | 0 | 0 | 11 | 100 |  |
| Evening | 0 | 0 | 2 | 18.18 | 8 | 72.72 | 1 | 9.10 | 11 | 100 |  |
| Total | 12 | 27.27 | 21 | 47.73 | 10 | 22.73 | 1 | 2.27 | 44 | 100 |  |

### Association between Sleep Quality and Work Fatigue in Phosphoric Acid Plant

Research results obtained using cross tabulation between sleep quality with work fatigue obtained the following data:

Based on table 5, it can be seen that the majority of respondents in the phosphoric acid plant with good sleep quality experienced low levels of fatigue by 78.57%, while respondents with poor sleep quality dominated moderate work fatigue by 63.34%. This indicates that respondents with poor sleep quality tend to experience high levels of fatigue compared to workers with good sleep quality. From the results of statistical analysis, it was found that the correlation coefficient between the age variable and the work fatigue variable was 0.619, which means the strength of the relationship between sleep quality and work fatigue is strong.

**Table 5.** Association of Work Fatigue based on Sleep Quality

| Sleep<br>Quality | Work Fatigue |  |  |  |  |  |  |  | Total |  | Correlation<br>Coefficient |
| --- | --- | --- | --- | --- | --- | --- | --- | --- | --- | --- | --- |
|  | Low |  | Average |  | High |  | Very<br>High |  |  |  |  |
|  | N | % | N | % | N | % | N | % | N | % |  |
| Good | 11 | 78.57 | 2 | 14.29 | 1 | 7.14 | 0 | 0 | 14 | 100 | 0.619 |
| Bad | 1 | 3.33 | 19 | 63.34 | 9 | 30.00 | 1 | 3.33 | 30 | 100 |  |
| Total | 12 | 27.27 | 21 | 47.73 | 10 | 22.73 | 1 | 2.27 | 44 | 100 |  |

## DISCUSSION

### Work Shift in Phosphoric Acid Plant

The results of this research related to the respondents’ work shifts are known that most of the respondents’ work on the morning shift as many as 22 respondents (50.0%). The number of respondents on the morning shift was 22 workers, while the afternoon shift and night shift each amounted to 11 people. This is because, data collection was carried out for two days. On the first day, data were collected on three work groups, namely the morning shift, afternoon shift, and night shift. Meanwhile, data collection for another working group was carried out on the second day, in which the working group worked on the morning shift.

Shift work is a work time that is run if two or more employees are a group working in the same or different time sequences (work rotation) and at the same workplace (Winarsunu, 2008). Shift work at this plant is divided into three work shifts, namely morning shift, afternoon shift, and night shift. The morning shift starts at seven in the morning until two in the afternoon, the afternoon shift starts at two in the afternoon until eleven at night, and then the night shift starts at eleven in the evening until seven in the morning. In the division of work shifts, workers are divided into four groups. In one day there will be three groups of workers working, while one group of workers will have a day off.

The work shift system at PT X, namely at the phosphoric acid plant, is that workers work two to three days in the morning shift, followed by two or three days in the afternoon shift. After working on the morning shift and afternoon shift, workers will get one day off, followed by working the night shift for two or three days. After working the night shift, workers will get one day off. Workers are also entitled to a day off on consecutive Saturdays and Sundays, once a month.

### Sleep Quality in Phosphoric Acid Plant

The results of the study related to the sleep quality of respondents, it is known that most of the respondents have poor sleep quality, namely as many as 30 respondents (68.18%). Measurement of sleep quality in this study used a Pittsburgh Sleep Quality Index (PQSI) questionnaire consisting of 18 questions. The questionnaire consisted of seven aspects of sleep quality including subjective sleep quality, sleep latency, sleep efficiency, use of sleeping pills, sleep disturbances, sleep duration, and disturbances in daily activities. There will be a decrease in sleep quality, if one of these aspects is disturbed. Sleep quality is said to be good if a person does not show signs of sleep deprivation and does not experience problems during sleep.

According to Safe Work Australia (2013) sleep quality can contribute and can increase the risk of fatigue. When a person experiences fatigue, the muscles can recover with rest, but the brain can only recover with sleep. The most beneficial sleep is a deep, uninterrupted sleep that is carried out in one continuous period. The optimal amount of sleep varies from person to person, but in general adults need seven to eight hours of sleep each day.

According to Kuswana (2017) one of the lifestyle factors related to fatigue is inadequate or poor sleep quality due to sleep disorders. When compared with this theory, respondents who mostly have poor sleep quality are at greater risk of experiencing fatigue.

### Work Fatigue in Phosphoric Acid Plant

The measurement of work fatigue in this study used a Subjective Feelings of Fatigue questionnaire from the Industrial Fatigue Research Committee (IFRC). The questionnaire has 30 questions consisting of 10 questions about the weakening of activities, 10 questions about the weakening of motivation, and 10 questions about the description of physical fatigue. The results of the research related to the work fatigue of the respondents, it is known that most of the respondents experienced moderate level of work fatigue, namely as many as 21 respondents (47.70%).

Fatigue is a protective mechanism of the body so that the body avoids further damage so that recovery occurs after rest. In general, the fatigue experienced by each person shows a different condition. In general, symptoms of fatigue start from very mild complaints to feelings of exhaustion (Tarwaka, 2014). A common symptom that is often felt in workers who experience fatigue is a loss of will to work. If the worker feels tremors or pain in the muscles, then the worker has experienced muscle fatigue (Suma’mur, 2014).

The causes of work fatigue in general are health conditions and nutritional conditions, an inappropriate work environment, disproportionate working and rest periods, and high physical or mental work intensity (Maurits, 2010). The influence of the conditions that cause fatigue such as gathering and piling up resulting in feelings of fatigue. High feelings of fatigue cause workers to no longer be able to work (Suma’mur, 2014).

One effort that can be done to reduce work fatigue is to rest and sleep. Adequate rest and sleep will allow the new body to function optimally. Rest means a state of calm, relaxation, without emotional stress, and free from feelings of restlessness. Resting does not mean not doing activities at all. Taking a walk in the park can sometimes also be regarded as a form of rest. Sleep is a state of altered consciousness when the individual’s perception and reaction to the environment decreases. Sleep is characterized by minimal physical activity, varying levels of consciousness, changes in the body’s physiological processes, and decreased response to external stimuli. Muscle fatigue can be recovered with rest, but the brain can only recover with sleep (Hidayat, 2012).

### Association between Work shift and Work Fatigue in Phosphoric Acid Plant

Based on the results of research that has been done, it can be seen that most of the workers on the morning shift experienced low levels of fatigue (45.45%) and moderate levels of fatigue (45.45%). Meanwhile, most of the afternoon shift workers experienced moderate fatigue (81.82). And the night shift workers, the majority experienced a high level of fatigue (72.72%). The correlation coefficient value is 0.637, which means that there is a strong relationship between work shifts and work fatigue.

This study is in line with research conducted by Hestya (2012) which states that there is a relationship between work shifts and fatigue levels. United Electrical (UE) News Health and Safety (1998) reported that long work shifts can result in indigestion, sleep disturbances, and fatigue. This research is also in line with research conducted by Pratama & Wijaya (2020) which states that there is a relationship between work shifts and fatigue with a p-value of 0.032.

The result of Relative Risk (RR) = 1.5 shows that night shift workers are 1.5 times more likely to experience fatigue than morning shift workers. In general, all body functions are in optimal condition for work is during the day. While at night is a time to rest and recover resources (energy). Disruption of these two phases causes disturbances that have an impact on a person’s physiology and psychology. All studied human functions exhibit regular daily cycles. Night shift work will have an impact on the body’s physiological response, social effects, and the effect of work appearance or performance (Pulat, 2002).

Based on the research results, respondents on the night shift tend to experience a high level of fatigue compared to respondents on the morning and evening shifts. This is in accordance with the theory of Wyatt and Mariot in Pulat (2002) which states that as a result of physiological and social effects, performance will also decrease at night.

Night work shifts need much attention because the rhythm of human physiology (circadian rhythm) is disturbed, the body’s metabolism cannot adapt, fatigue, lack of sleep, digestive organs are not functioning normally, psychological reactions and cumulative effects arise (Suma’mur, 2009). Circadian conditions that are disrupted at night are also the cause of fatigue in workers because body functions are not appropriate where the body is active at night and rests during the day. The circadian rhythm explains that for 24 hours the body has 2 phases, namely the ergotrophic phase where during the day all organs and body functions are ready to take an action, and the trophic phase where at night the body renews its energy reserves or re-strengthens (Winarsunu, 2008).

### Association between Sleep Quality and Work Fatigue in Phosphoric Acid Plant

Based on the results of the research that has been done, it can be seen that most of the workers have poor sleep quality, as many as 30 workers with a percentage of 63.34% experiencing moderate level of work fatigue. The results also show that the correlation coefficient value is 0.619, which means that there is a strong relationship between sleep quality and work fatigue.

The results of this study are in line with research conducted by Utami (2008) regarding the relationship between individual characteristics, work attitudes, and sleep quality with work fatigue in hospital nurses which shows that there is a relationship between sleep quality and work fatigue. Research conducted by Pratama & Wijaya (2020) also shows that there is a relationship between sleep quality and work fatigue in workers who live in the mess of PT. Pamapersada jobsite MTBU South Sumatra. The results of this study indicate a Relative Risk (RR) of 1.411, meaning that workers with poor sleep quality are 1.4 times more likely to experience high levels of fatigue when compared to workers with good sleep quality.

In general, humans work in the morning and during the day, while at night to rest and sleep. Generally all bodily functions increase during the day, begin to weaken in the afternoon, decline at night for recovery and renewal. Things like this follow a biological clock pattern called circadian rhythms (Pulat, 1992).

Based on the results of the study, most workers have poor sleep quality, this can be caused by the lack of rest time they have. While working, workers cannot use the rest time provided by the company to sleep. When break time comes, they use it to eat and chat with other workers. When workers get one to two days off after the end of a work shift, they are sometimes asked to come in to replace workers from other groups who are not in. This can lead to a lack of rest time for workers at home due to, among other factors, the home environment, for example not being able to sleep because the baby is crying, playing with children, staying up late watching television, and the lack of rest time to sleep at night because they have to work. until the evening.

The habit of short duration is also one of the factors that cause poor sleep quality. Physical and cognitive symptoms of poor sleep quality include fatigue, loss of concentration, pain, anxiety, restlessness, irrational thoughts, hallucinations, loss of appetite, constipation, and proneness to accidents (Buysee, 1998).

## CONCLUSION

Based on the results of data processing using descriptive statistical analysis, relationship analysis using the Spearman test and contingency coefficient, and the discussions that have been carried out previously, the following conclusions can be drawn:

1. The sleep quality of workers who are dominated by phosphoric acid plant workers is poor sleep quality.
2. The most common work fatigue experienced by workers at the phosphoric acid plant is moderate work fatigue.
3. The relationship between work shifts and fatigue is a strong relationship.
4. The relationship between sleep quality and fatigue is strong.
5. Special inspection at 2-4 am, because it is a crucial time, which many workers will be sleepy and increase work accident risk

## Supporting information

Ethical Acceptance

## Data Availability

All data produced in the present study are available upon reasonable request to the authors

