## Supplementary material for "Work Fatigue in Phosphoric Acid Industry Workers: How Work shift and Sleep Quality Affect Them?": Ethical Acceptance

#### UNIVERSITAS AIRLANGGA FACULTY OF DENTAL MEDICINE
#### HEALTH RESEARCH ETHICAL CLEARANCE COMMISSION

### ETHICAL CLEARANCE CERTIFICATE

**Number : 275/HRECC.FODM/VI/2021**

Universitas Airlangga Faculty Of Dental Medicine Health Research Ethical Clearance Commission has studied the proposed research design carefully, Declared to be ethically appropriate in accordance to 7 (seven) WHO 2011, and therefore, shall herewith certify that the research entitled :

**"Relationship between Work Shift and Sleep Quality with Subjective Work Fatigue on Production Section Workers of PT. X"**

Principal Researcher : ANGELA TESALONIKA OKTAVERA

Unit/Institution/Place of Research : - PT. X

**CERTIFIED TO BE ETHICALLY CLEARED**

PANITIA KELAIKAN ETIK FAKULTAS KEDOKTERAN GIGI UNIVERSITAS AIRLANGGA

Surabaya, May 31, 2021
Chairman,

Prof.Dr. TAMARA YUANITA, drg.,MS.,Sp.KG(K)
Official No. 196006251986012002
